# Is Human Bocavirus Infection Associated with Gastroenteritis in Children? An Updated Systematic Review and Meta-analysis

**DOI:** 10.1101/2021.10.08.21264786

**Authors:** Minyi Zhang, Minyi Liang, Qiushuang Li, Juxian Xian, Fei Wu, Liting Zheng, Qing Chen

## Abstract

**Background:** Human bocavirus (HBoV) figures as an increased risk factor of respiratory and gastrointestinal tract infections among children. A great deal of data is available to support the pathogenic role of HBoV in acute respiratory diseases. However, the association between HBoV infection and gastroenteritis remains controversial due to the ambiguous results. The present work aims to clarify the role of HBoV as a cause of gastroenteritis in children.

**Methodology/Principal findings:** A systematic search of the literature was carried out from 1 January 2016 to 29 August 2021 in Embase, PubMed, Scopus, Web of Science, and the Chinese bibliographic database of biomedicine (CBM). Data from included studies were analyzed by use of a random-effects model. The pooled estimates of HBoV prevalence among all cases of gastroenteritis were generated and stratified by potential confounders. The pooled odds ratio (OR) and 95% confidence interval (CI) were computed for HBoV infection in relation to the risk of gastroenteritis. The overall prevalence of HBoV in children with gastroenteritis (9.1%, 95% CI: 6.7-11.8%) was considerably higher than that detected in children without gastroenteritis (4.0%, 95% CI: 1.1-8.5%). HBoV prevalence tended to be higher in cases of gastroenteritis under five years of age (12.1%, 95% CI: 6.8-18.7%). The highest frequency of HBoV was found in Egypt (57.8%, 95% CI: 47.7-67.6%). The predominant genotypes of HBoV circulating in children with gastroenteritis were genotype 1 (HBoV1, 3.8%, 95% CI: 2.7-5.2%) and genotype 2 (HBoV2, 2.4%, 95% CI: 1.3-3.7%). HBoV infection was significantly associated with an increased risk of gastroenteritis in children (OR 1.620, 95% CI: 1.023-2.566).

**Conclusion:** The HBoV prevalence in pediatric cases of gastroenteritis is higher than that in children without gastroenteritis, demonstrating an increasing global burden of gastroenteritis in children caused by HBoV infection. Targeted intervention to reduce the HBoV burden should be established.

**Author summary:** In addition to the known gastroenteritis-associated pathogens (rotavirus and norovirus et al.), several novel viruses that might be caused agents of diarrhea have been gradually determined, such as HBoV. The impact of HBoV infection on the development of childhood gastroenteritis remains ambiguous and in need of verifications. This work clarifies the association between HBoV infection and the risk of gastroenteritis in children based on the review literature. The estimated HBoV prevalence in all cases of gastroenteritis was categorized into different subgroups following the potential confounders, including age, gender, country, and HBoV genotypes. The main finding of this work was the significant association between HBoV infection and the development of childhood gastroenteritis. Our systematic review highlighted that HBoV infection contributes to the increased global burden of gastroenteritis in children.

## Introduction

Childhood Diarrheal disease is a leading cause of malnutrition and death in children younger than five years, which accounted for roughly 1.7 billion novel cases and greater than 500 thousand deaths each year throughout the world (1). Infection diarrhea is one of the most frequent clinical manifestations of gastrointestinal tract infection resulting from either bacteria, viruses, parasites, or fungi (2). As being preventable, fully understanding its causative agents would be of vital benefit from substantially decreasing the burden of diarrheal disease in children. Of these, viruses turn out to be the primary causative agents responsible for around 70% of gastroenteritis cases in children (3, 4). For decades, rotavirus was the primary pathogen of gastroenteritis-related hospitalizations and other severe diseases in the first five years of life (5). However, the recent wide application of rotavirus vaccines is an essential contributor to the marked decrease in rotavirus-related illnesses (6, 7). Since then, other novel detected viral pathogens have become the potential risk factors of childhood gastroenteritis, such as human bocavirus (HBoV).

HBoV consists of ∼5kb linear and small non-enveloped single-stranded DNA genome organizing in three open reading frames that encode two forms of non-structural protein 1 (NS1), a nuclear phosphoprotein (NP1), and two viral capsid proteins (VP1 and VP2) (8-10). Following the genomic analysis, HBoV belongs to the genus *Bocavirus* within the family *Parvoviridae* and the subfamily *Parvovirinae*, with four currently identified genotypes. HBoV genotype 1 (HBoV1) was firstly discovered in nasopharyngeal aspirates from children with respiratory tract infections (5). With its initial discovery in 2005 by Tobias Allander and colleagues (5), numerous studies regarding the epidemiology, pathogenesis, diagnosis, and clinical features of this virus have been carried out in many countries. Subsequently, three additional genotypes were detected in faces and named HBoV2, 3, and 4 (11, 12). A great deal of data is available to support the association between HBoV1 infection and acute respiratory diseases, such as asthma, pneumonia, bronchiolitis, and acute wheezing (13-17). On the contrary, the remaining genotypes are primarily identified in human specimens. Previous studies have suggested a comparable prevalence of HBoV in patients with gastroenteritis compared to healthy or non-diarrheal individuals (11, 18). For instance, a prior study involving 632 children with gastroenteritis and 162 healthy controls has shown a statistically significant association between HBoV infection and childhood gastroenteritis (18). It should be noted that, in the same study, the majority of HBoV infections were found in children less than 12 months of age (18). Most recently, Neveen et al. concluded that HBoV could be the significant etiologic agent of gastroenteritis due to the presence of HBoV in some children suffering from diarrheal disease without any association of any other etiological agents (19). However, a controversial issue that arises in this domain is the actual pathogen role of HBoV in gastrointestinal symptoms due to the growing appears of HBoV detection in asymptomatic patients (20, 21).

A systematic review and meta-analysis have been published to assess whether HBoV infection is a risk factor for gastroenteritis in children under five years (22). This review included 36 unique studies from 18 countries between Oct. 2005 and Oct. 2016 and estimated that 6.90% of all cases of gastroenteritis in children younger than five years were correlated with HBoV infection. However, this review failed to identify a statistically significant association of HBoV infection with the risk of gastroenteritis. Also, the prevalence and distribution of HBoV3 and HBoV4 are needed to be established due to the lack of pooled estimations.

With the increasing use of molecular diagnostics and much attention of scientists, the data on different genotypes of HBoV associated with gastroenteritis in children has increased in the last five years and accordingly prompted us to update previous estimates of HBoV prevalence and genotype distribution in pediatric cases of gastroenteritis up to 29 August 2021. Additionally, the second aim of this work was to determine the association between HBoV infection and gastroenteritis in children through a meta-analytic method.

## Methods

### Data sources

Following the Preferred Reporting Items for Systematic Review and Meta-Analyses (PRISMA) statement (23), a comprehensive literature search was conducted in Embase, PubMed, Scopus, Web of Science, and CBM (Chinese bibliographic database of biomedicine) databases for identifying eligible records from 1 January 2016 to 29 August 2021. The Medical Subject Heading (MeSH) terms “human bocavirus”, “gastroenteritis”, “diarrhea”, “child”, and related terms were used in all possible combinations. More detailed search strategies in each database are showed in S1 Table. Since fewer studies in a case-control design were identified during the present time horizon of the literature search, we additionally included the case-control studies from a previous systematic by Ri De and colleagues (22) and extracted data for other variables as required.

### Study selection

Studies that met the following criteria were deemed eligible for inclusion: 1) enrolled pediatric patients with gastroenteritis or healthy children; 2) applied PCR-based diagnostics for HBoV on fecal samples; 3) original cross-sectional or case-control documents of HBoV infection. The exclusion criteria were the following: 1) duplicate records; 2) HBoV prevalence in adults; 3) samples other than stools; 4) HBoV infection employing antigen assays and studies that reported seroprevalence of HBoV antibodies; 5) lack of detailed data; 6) animal studies, conference abstracts, reviews, letters, case reports, or obviously irrelevant.

### Data extraction

Two researchers independently reviewed and selected all potentially relevant articles following the inclusion and exclusion criteria for further assessment. Suppose any conflicts exist between the investigators, a third evaluator will make a final decision. The following information was extracted from the included publications by two researchers: first author, year of publication, country, study period, study design, used diagnostic method, age range or mean, gender, sample size, number of total cases, number of HBoV-positive cases, number of total controls, number of HBoV-positive controls, and genotypes of HBoV.

### Assessment of Study Quality

Two investigators independently assessed the quality of each retrieved study in a case-control design through the Newcastle-Ottawa Scale (NOS) (24). A study was awarded a maximum of one star for each numbered item within the selection and exposure sections. A maximum of two stars was awarded regarding the comparability category. Studies that received seven or more out of a maximum of 9 stars were judged as being of high quality. Meanwhile, medium-quality and poor-quality studies were considered those with four to six stars and those with fewer than four stars, respectively.

### Statistical analysis

Using the HBoV prevalence as the primary outcome, we firstly generated pooled estimated prevalence of HBoV infection in all pediatric patients with gastroenteritis. Further, a meta-analysis of risk effect estimates was carried out regarding the HBoV exposure and gastroenteritis in children by the DerSimonian and Laird random-effects model (25). Pooled estimates of odds ratios (ORs) and 95% confidence interval (CIs) were computed. The *I*^*2*^ statistic was utilized to measure the proportion of the total variability explained by heterogeneity across studies. *I*^*2*^ values < 25% indicate a low level of heterogeneity, whereas *I*^*2*^ values ≥ 75% is regarded as a high degree of heterogeneity (26). Cochrane’s *Q* statistic was applied to examine the statistical significance of the heterogeneity. We stratified the pediatric patients based on age, gender, country, and HBoV genotype and subsequently calculated the summary risk of gastroenteritis for HBoV infection. A Pearson’s chi-square test was employed to identify the significant difference between the subgroups. Additionally, we addressed whether the distribution of HBoV infection varied between pediatric cases of gastroenteritis and healthy children.

Publication bias was assessed by combining the funnel plot, Begg’s adjusted rank correlation test, and Egger’s regression asymmetry test (27, 28). All statistical analyses were conducted in Stata 16. A probability level less than 0.05 was considered statistically significant except the heterogeneity test set at less than 0.1, and all statistical tests were two-sided.

## Results

### Literature search

Based on the search strategy, seven hundred and seventy-three relevant articles were initially retrieved from Embase, PubMed, Scopus, Web of Science, and CBM, of which 266 were finally identified between 2016 and 2021. One hundred and twenty-seven documents were subsequently removed due to duplication, and a total of 222 records were excluded after manual screening of titles and abstracts. The remaining 44 full-text articles were further obtained for eligibility, of which 29 unique studies were included after applying the inclusion and exclusion criteria in this systematic review and meta-analysis. Case-control studies originally identified by Ri De and colleagues were appended to our search. However, two of the ten studies were excluded due to reporting HBoV prevalence either in serum samples or in adults. Also, one study not published in original research was removed, leaving seven additional studies in a case-control design. Consequently, the present updated review consisted of 36 studies. A PRISMA-based flowchart of the literature search and study selection is depicted in Fig 1.

**Fig 1.**
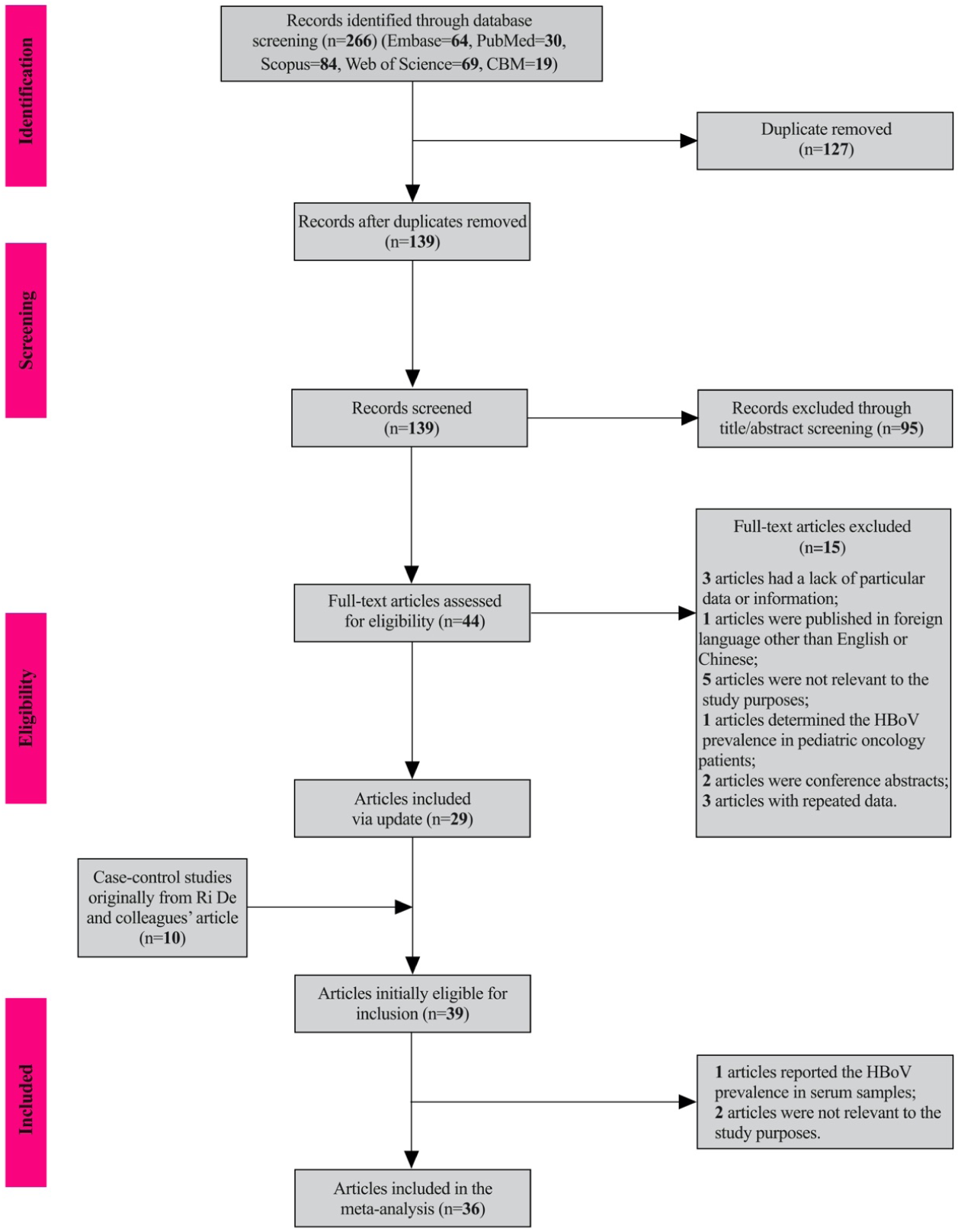
Flowchart of the literature search and study selection.

### Study characteristics

The final dataset included data on children from 20 countries (Russia, United States, India, China, Brazil, Albania, Korea, Japan, Sudan, Gabon, Latvia, Iraq, Iran, South Africa, Bangladesh, Australia, Finland, United Kingdom, Thailand, and Egypt). Of the 36 included documents, 11 were case-control (11, 18, 29-37) and 25 were cross-sectional in study design (20, 38-61). Amidst the cross-sectional studies, 23 were case-only, and the remaining 2 presented data for healthy individuals. The largest study investigated 5250 pediatric patients with gastroenteritis in Russia, and the smallest one comprised 83 cases. This review consisted of 15 and 29 studies reporting gender and genotype distribution of HBoV infection, respectively. The majority of these data represented children less than five years of age (n=20). Most studies were carried out in China (n=8), followed by Brazil (n=4) and Bangladesh (n=3). The characteristics of cross-sectional and case-control studies are summarized in Table 1 and 2.

**Table 1.**
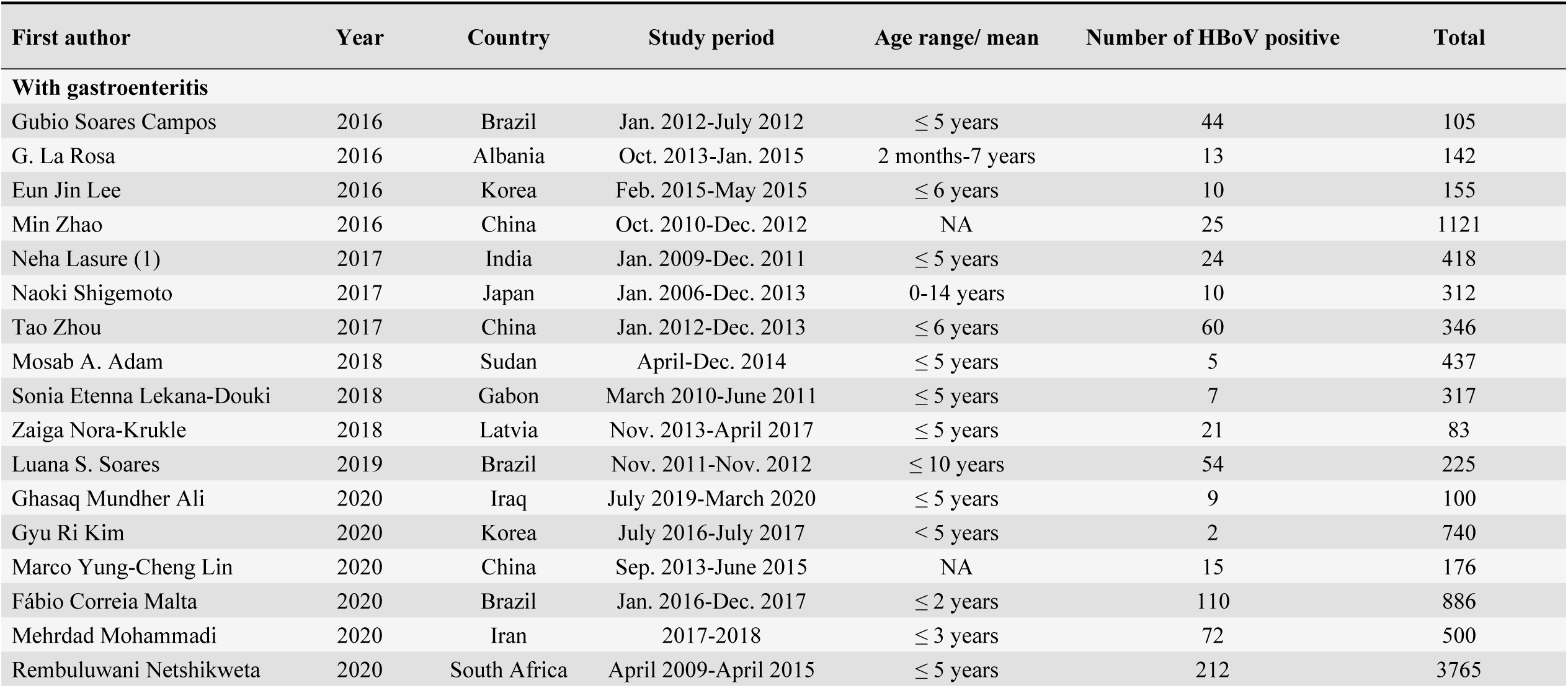
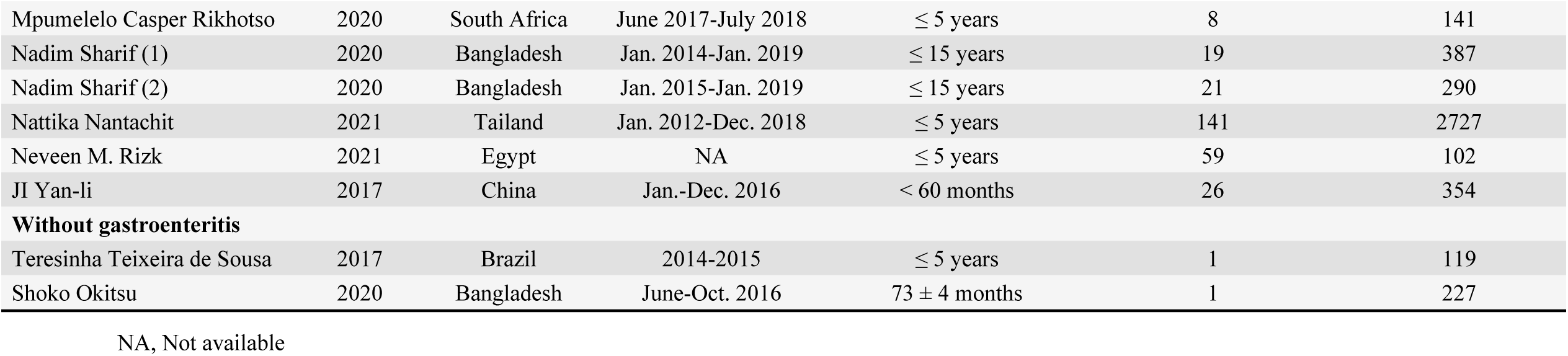
Summary of characteristics of cross-sectional studies.

| First author | Year | Country | Study period | Age range/ mean | Number of HBoV positive | Total |
| --- | --- | --- | --- | --- | --- | --- |
| **With gastroenteritis** |  |  |  |  |  |  |
| Gubio Soares Campos | 2016 | Brazil | Jan. 2012-July 2012 | ≤ 5 years | 44 | 105 |
| G. La Rosa | 2016 | Albania | Oct. 2013-Jan. 2015 | 2 months-7 years | 13 | 142 |
| Eun Jin Lee | 2016 | Korea | Feb. 2015-May 2015 | ≤ 6 years | 10 | 155 |
| Min Zhao | 2016 | China | Oct. 2010-Dec. 2012 | NA | 25 | 1121 |
| Neha Lasure (1) | 2017 | India | Jan. 2009-Dec. 2011 | ≤ 5 years | 24 | 418 |
| Naoki Shigemoto | 2017 | Japan | Jan. 2006-Dec. 2013 | 0-14 years | 10 | 312 |
| Tao Zhou | 2017 | China | Jan. 2012-Dec. 2013 | ≤ 6 years | 60 | 346 |
| Mosab A. Adam | 2018 | Sudan | April-Dec. 2014 | ≤ 5 years | 5 | 437 |
| Sonia Etenna Lekana-Douki | 2018 | Gabon | March 2010-June 2011 | ≤ 5 years | 7 | 317 |
| Zaiga Nora-Krukle | 2018 | Latvia | Nov. 2013-April 2017 | ≤ 5 years | 21 | 83 |
| Luana S. Soares | 2019 | Brazil | Nov. 2011-Nov. 2012 | ≤ 10 years | 54 | 225 |
| Ghasaq Mundher Ali | 2020 | Iraq | July 2019-March 2020 | ≤ 5 years | 9 | 100 |
| Gyu Ri Kim | 2020 | Korea | July 2016-July 2017 | < 5 years | 2 | 740 |
| Marco Yung-Cheng Lin | 2020 | China | Sep. 2013-June 2015 | NA | 15 | 176 |
| Fábio Correia Malta | 2020 | Brazil | Jan. 2016-Dec. 2017 | ≤ 2 years | 110 | 886 |
| Mehrdad Mohammadi | 2020 | Iran | 2017-2018 | ≤ 3 years | 72 | 500 |
| Rembuluwani Netshikweta | 2020 | South Africa | April 2009-April 2015 | ≤ 5 years | 212 | 3765 |
| Mpumelelo Casper Rikhotso | 2020 | South Africa | June 2017-July 2018 | $\leq 5$ years | 8 | 141 |
| Nadim Sharif (1) | 2020 | Bangladesh | Jan. 2014-Jan. 2019 | $\leq 15$ years | 19 | 387 |
| Nadim Sharif (2) | 2020 | Bangladesh | Jan. 2015-Jan. 2019 | $\leq 15$ years | 21 | 290 |
| Nattika Nantachit | 2021 | Tailand | Jan. 2012-Dec. 2018 | $\leq 5$ years | 141 | 2727 |
| Neveen M. Rizk | 2021 | Egypt | NA | $\leq 5$ years | 59 | 102 |
| JI Yan-li | 2017 | China | Jan.-Dec. 2016 | $< 60$ months | 26 | 354 |
| **Without gastroenteritis** |  |  |  |  |  |  |
| Teresinha Teixeira de Sousa | 2017 | Brazil | 2014-2015 | $\leq 5$ years | 1 | 119 |
| Shoko Okitsu | 2020 | Bangladesh | June-Oct. 2016 | $73 \pm 4$ months | 1 | 227 |

NA, Not available

**Table 2.** Summary of case-control studies.

| First author | Year | Country | Study period | Sample size | Age range/<br>mean | Number of<br>HBoV positive | Total | Age range/<br>mean | Number of<br>HBoV positive | Total |
| --- | --- | --- | --- | --- | --- | --- | --- | --- | --- | --- |
| Alexander Tymentsev | 2016 | Russia | Feb 2010-Dec 2012 | 5502 | ≤ 5 years | 62 | 5250 | ≤ 5 years | 1 | 252 |
| Rimma Melamed | 2017 | Unite States | Nov 2003-Nov 2005 | 356 | 44.8 (11-60)<br>months | 22 | 178 | 43.8 (11-59)<br>months | 14 | 178 |
| Neha Lasure (2) | 2019 | India | 2007-2011 | 985 | ≤ 5 years | 46 | 778 | ≤ 5 years | 0 | 207 |
| Ri De | 2018 | China | May-Dec 2016 | 480 | ≤ 5 years | 18 | 240 | ≤ 5 years | 12 | 240 |
| Jane L. Arthur<sup>a</sup> | 2009 | Australia | Jan-Dec 2001 | 372 | ≤ 18 years | 54 | 186 | ≤ 18 years | 30 | 186 |
| Minna Risku<sup>a</sup> | 2012 | Finland | Aug 2006- Aug<br>2008 | 990 | ≤ 15 years | 85 | 878 | ≤ 15 years | 6 | 112 |
| Sameena Nawaz<sup>a</sup> | 2012 | United Kingdom | 1993-1996 | 1772 | ≤ 19 years | 106 | 851 | ≤ 19 years | 163 | 921 |
| Thaweesak Chieochansin<sup>a</sup> | 2008 | Thailand | Nov 2005- Sep 2007 | 427 | 4 months-4<br>years | 2 | 225 | 2 months-5<br>years | 0 | 202 |
| Wei-xia Cheng<sup>a</sup> | 2008 | China | July 2006-Sep 2007 | 512 | ≤ 5 years | 14 | 397 | ≤ 5 years | 4 | 115 |
| Yu Jin<sup>a</sup> | 2011 | China | July 2006- June<br>2008 | 794 | ≤ 5 years | 162 | 632 | ≤ 5 years | 24 | 162 |
| Zeng Mei<sup>a</sup> | 2010 | China | Oct-Dec 2008 | 420 | ≤ 5 years | 6 | 220 | ≤ 5 years | 4 | 200 |

### HBoV prevalence in children with gastroenteritis

The overall pooled prevalence of HBoV infection among 23,664 pediatric cases of gastroenteritis was estimated at 9.1% (95% CI: 6.7-11.8%; *I*^*2*^ = 97.7%, *P* < 0.001 for heterogeneity test) (Fig 2). Subgroup analysis demonstrated the prevalence of HBoV exposure significantly differed across age category, country, and genotype (Table 3). By age, HBoV prevalence was significantly higher in cases of gastroenteritis younger than 5 years (12.1%, 95% CI: 6.8-18.7%), and the lowest prevalence was found in children between 5 and 10 years (3.4%, 95% CI: 0.2-8.6) (*P* < 0.001). By country, the highest prevalence of HBoV infection occurred in Egypt at 57.8% (95% CI: 47.7-67.6%), followed by Australia (29.0%, 95% CI: 22.6-36.1%) and Latvia (25.3%, 95% CI: 16.4-36.0%), and the lowest one was observed in Korea (0.7%, 95% CI: 0.2-1.4%) (*P* < 0.001). HBoV prevalence was slightly greater in male patients (9.4%, 95% CI: 5.8-13.8%) compared with female patients (8.7%, 95% CI: 5.7-12.2%) (*P* = 0.785). The estimated pooled HBoV prevalence did not markedly change when meta-analysis was limited to the studies that lasted at least 1 year (8.1%, 95% CI: 5.7-10.8, *P* = 0.221, *I*^*2*^ = 97.7%, *P* < 0.001 for heterogeneity test, n=27) (S1 Fig).

**Table 3.** Estimated HBoV prevalence in children with gastroenteritis by age, gender, country, and genotype.

| Category | Subgroup | Number of studies | Pooled prevalence (% , 95% CI) | Heterogeneity test (I<sup>2</sup>%, *P* value) | Chi-square test (*P* value) |
| --- | --- | --- | --- | --- | --- |
| **Age** | ≤ 1 year | 8 | 7.3 (3.6-12.2) | 97.142%, < 0.001 | *P* < 0.001 |
|  | > 1-5 years | 10 | 5.2 (1.8-9.9) | 96.373%, < 0.001 |  |
|  | ≤ 5 years | 13 | 12.1 (6.8-18.7) | 97.921%, < 0.001 |  |
|  | > 5-10 years | 2 | 3.4 (0.2-8.6) | < 0.001, < 0.001 |  |
|  | Mixed | 12 | 8.3 (6.2-10.7) | 96.949%, < 0.001 |  |
| **Genger** | Male | 14 | 9.4 (5.8-13.8) | 95.789%, < 0.001 | *P* = 0.785 |
|  | Female | 14 | 8.7 (5.7-12.2) | 92.220%, < 0.001 |  |
| **Country** | Russia | 1 | 1.2 (0.9-1.5) | NA | *P* < 0.001 |
|  | Unite States | 1 | 12.4 (7.9-18.1) | NA |  |
|  | India | 2 | 5.8 (4.6-7.3) | < 0.001, <0.001 |  |
|  | China | 8 | 8.2 (3.1-15.2) | 97.572%, <0.001 |  |
|  | Brazil | 3 | 24.7 (10.8-42.0) | < 0.001, <0.001 |  |
|  | Albania | 1 | 9.2 (5.0-15.1) | NA |  |
|  | Korea | 2 | 0.7 (0.2-1.4) | < 0.001, <0.001 |  |
|  | Japan | 1 | 3.2 (1.5-5.8) | NA |  |
|  | Sudan | 1 | 1.1 (0.4-2.6) | NA |  |
|  | Gabon | 1 | 2.2 (0.9-4.5) | NA |  |
|  | Latvia | 1 | 25.3 (16.4-36.0) | NA |  |
|  | Iraq | 1 | 9.0 (4.2-16.4) | NA |  |
|  | Iran | 1 | 14.4 (11.4-17.8) | NA |  |
|  | South Africa | 2 | 5.5 (4.8-6.2) | < 0.001, <0.001 |  |
|  | Bangladesh | 2 | 5.9 (4.2-7.8) | < 0.001, <0.001 |  |
|  | Australia | 1 | 29.0 (22.6-36.1) | NA |  |
|  | Finland | 1 | 9.7 (7.8-11.8) | NA |  |
|  | United Kingdom | 1 | 12.5 (10.3-14.9) | NA |  |
|  | Tailand | 2 | 4.7 (3.9-5.5) | < 0.001, <0.001 |  |
|  | Egypt | 1 | 57.8 (47.7-67.6) | NA |  |
| **Genotype** | HBoV1 | 28 | 3.8 (2.7-5.2) | 94.878%, < 0.001 | *P* < 0.001 |
|  | HBoV2 | 24 | 2.4 (1.3-3.7) | 96.249%, < 0.001 |  |
|  | HBoV3 | 21 | 0.9 (0.4-1.6) | 93.467%, < 0.001 |  |
|  | HBoV4 | 16 | 0 | 14.799, 0.284 |  |

**Fig 2.**
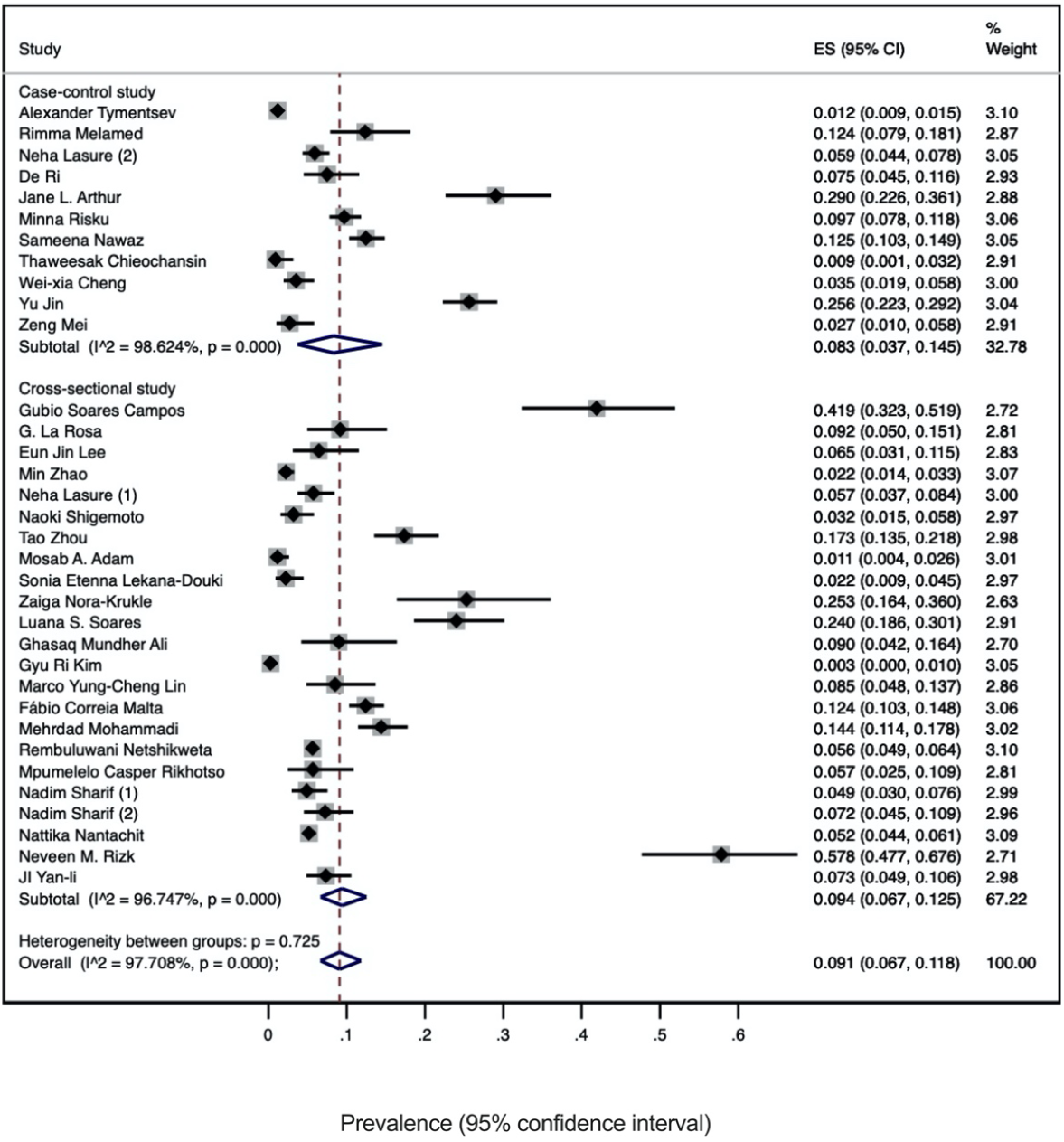
Overall pooled prevalence of HBoV infection in children with gastroenteritis based on case-control and cross-sectional studies. HBoV, human bocavirus

Whether the distribution of HBoV infection differed across pediatric cases of gastroenteritis and healthy children was further explored. In the 13 studies that included 3121 children without gastroenteritis, the pooled HBoV prevalence was detected in 4.0% (95% CI: 1.1-8.5%) (S2 Fig), which was significantly lower than that in cases of gastroenteritis (*P* < 0.001).

### Prevalence and distribution of HBoV genotypes

All four genotypes of HBoV were identified among pediatric cases of gastroenteritis (Table 3). The predominant genotype of HBoV circulating in children with gastroenteritis belonged to genotype 1 (HBoV1, 3.8%, 95% CI: 2.7-5.2%) and genotype 2 (HBoV2, 2.4%, 95% CI: 1.3-3.7%). Sixteen countries had reported the genotype distribution of HBoV. At the global level, HBoV1 dominated (50.5%), followed by HBoV2 (36.6%) and HBoV3 (12.0%). The estimated genotype distribution by country is depicted in Fig 3.

**Fig 3.**
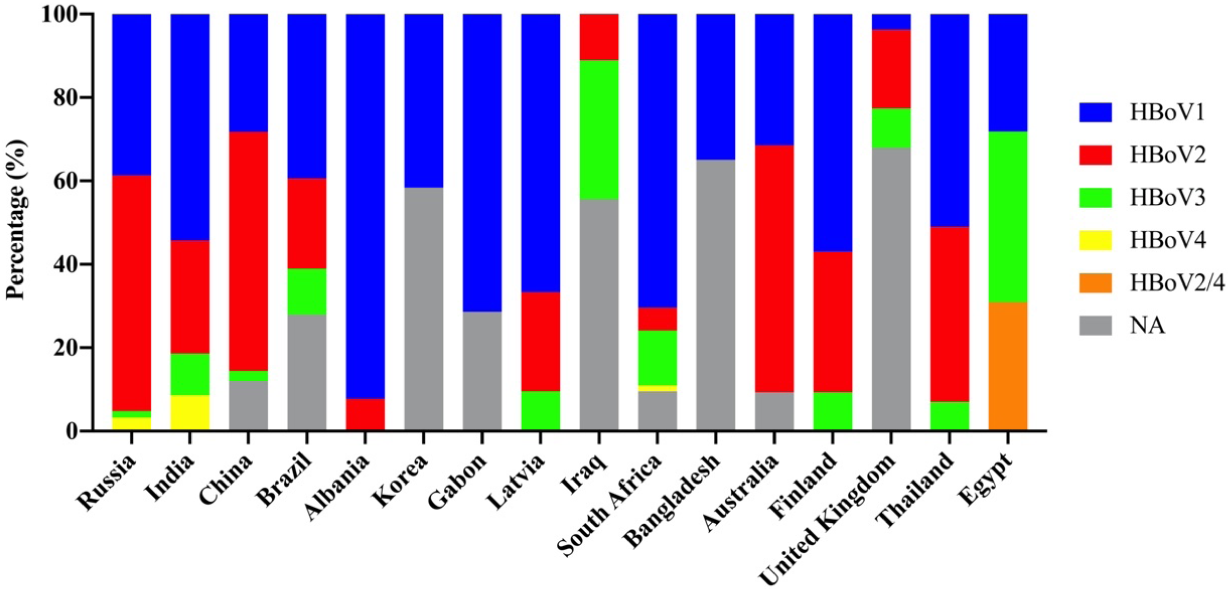
Genotype distribution by country.

### Association between HBoV infection and the risk of gastroenteritis in children

A quality assessment was performed for each study with a case-control design utilizing the NOS recommended by the Cochrane Collaboration (62). All eleven included case-control studies met the criteria for NOS and none of the poor quality (S2 Table). We conducted the second analysis based on case-control studies to investigate the relationship between HBoV infection and the risk of gastroenteritis in children, with the pooled OR value of 1.620 (95% CI: 1.023-2.566; *I*^*2*^ = 73.6%, *P* < 0.001 for heterogeneity test) being reported (Fig 4). In regard to HBoV genotypes, only HBoV2 showed a statistically significant correlation with gastroenteritis in children (OR 1.75, 95% CI: 1.30-2.35). Among different countries, the significant association of HBoV infection with gastroenteritis was observed in India, China, and Australia. Detailed associations between HBoV infection and gastroenteritis for subgroups were displayed in Fig 5. Using a visual inspection of the funnel plot and statistical tests, no evidence of publication bias was recognized (*P* = 0.876 for Begg’s test and *P* = 0.145 for Egger’s test) (Fig 6).

**Fig 4.**
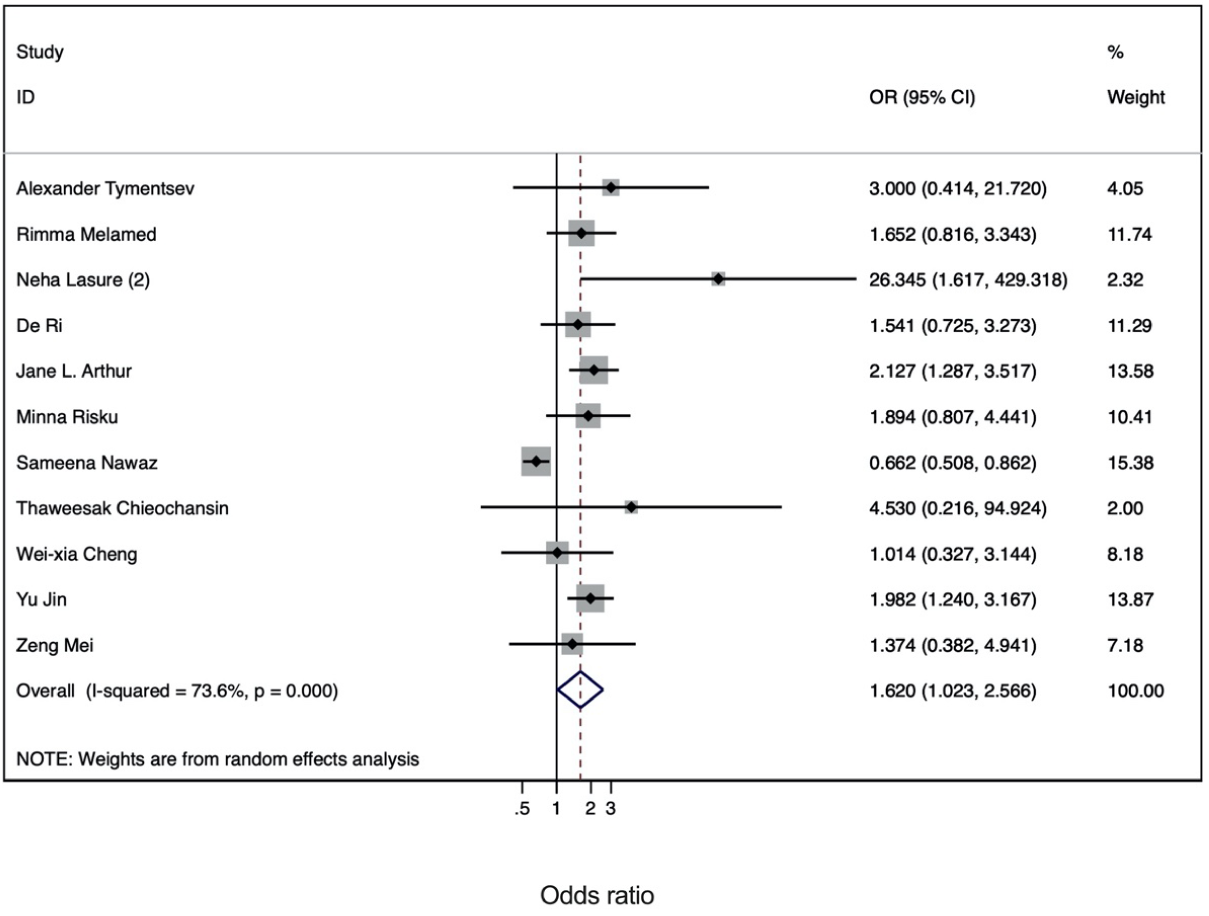
Forest plot of the association between HBoV infection and the risk of gastroenteritis in children. HBoV, human bocavirus; OR, odds ratio; CI, confidence interval

**Fig 5.**
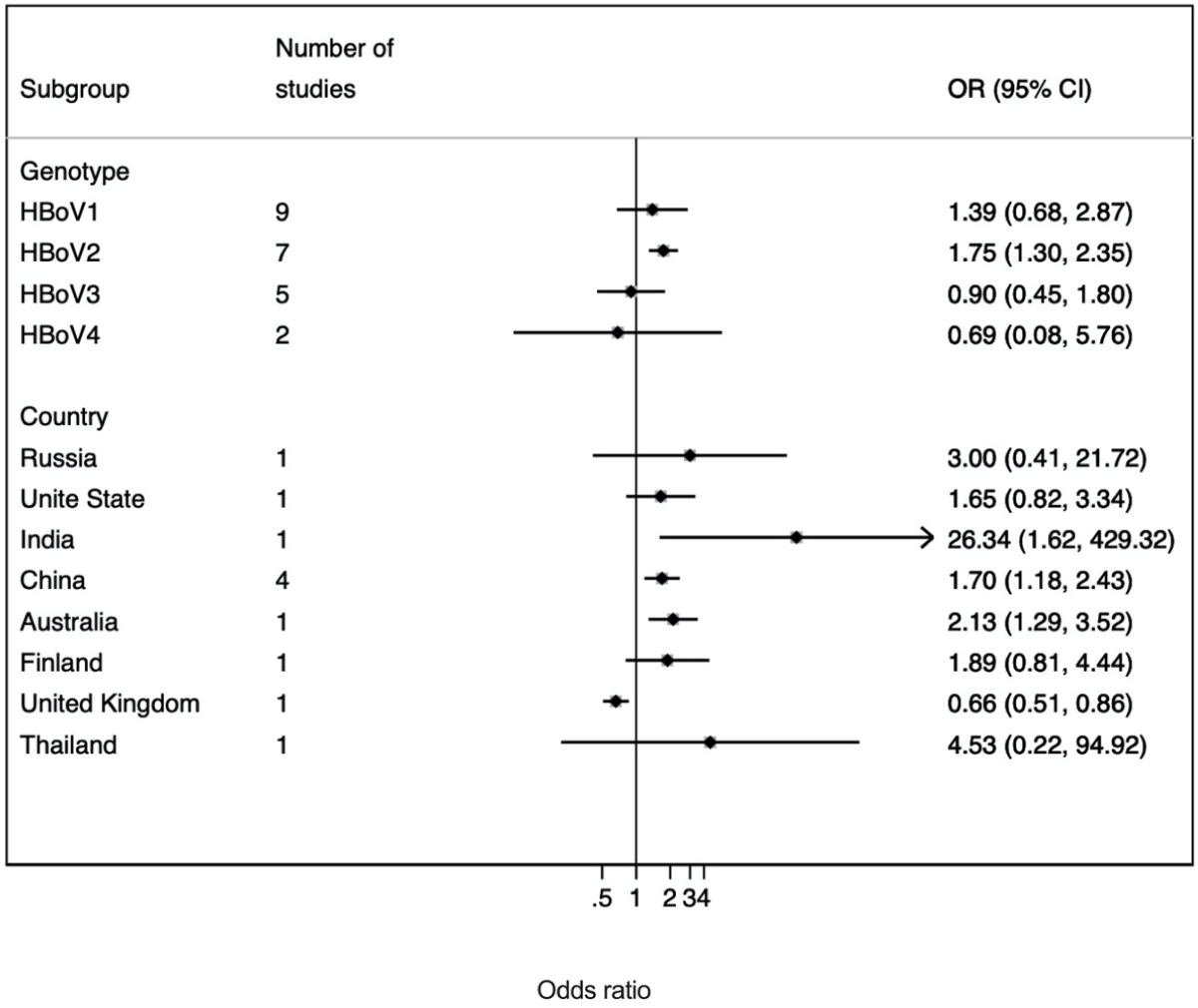
Forest plot of the association between HBoV infection and the risk of gastroenteritis in children by genotype and country. HBoV, human bocavirus; OR, odds ratio; CI, confidence interval

**Fig 6.**
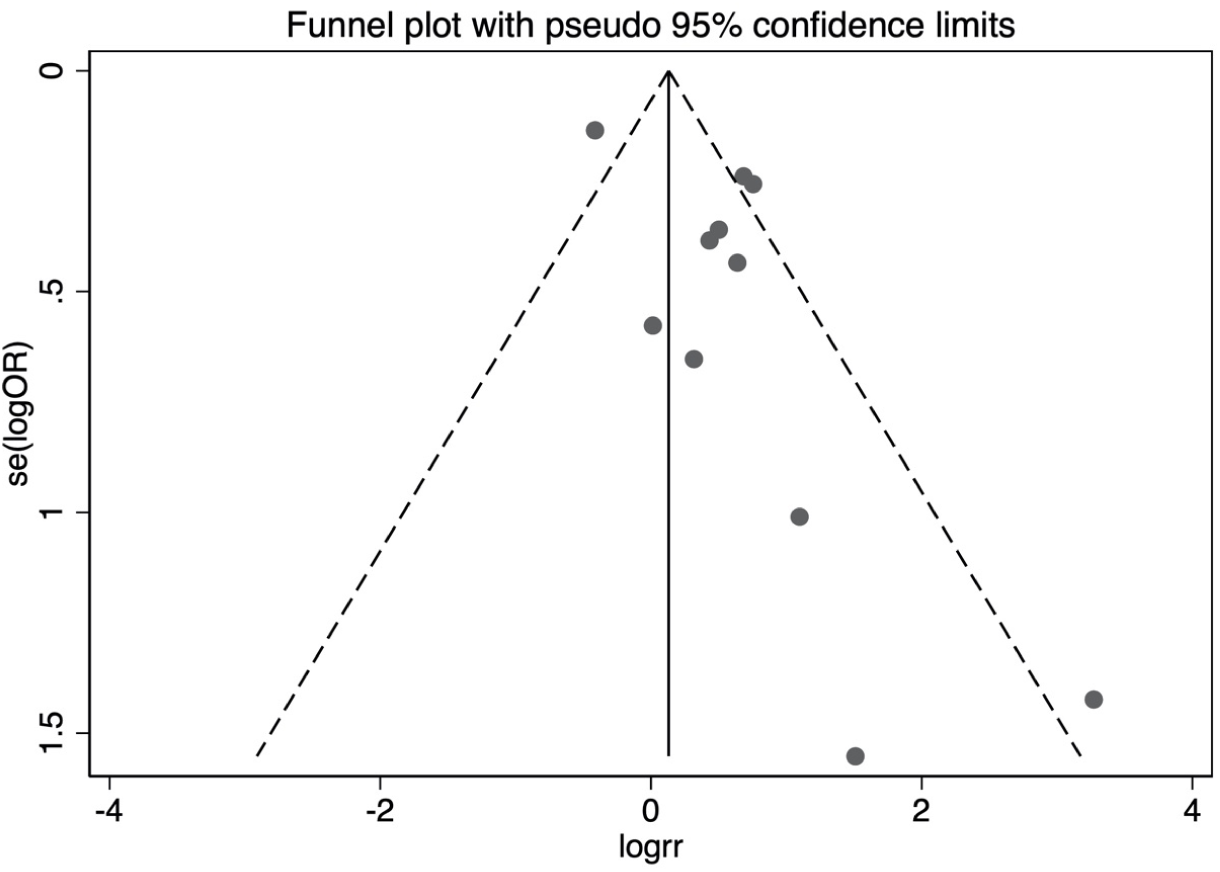
Funnel plot for assessment of publication bias.

## Discussion

HBoV has been considered a major pathogen that contributed to respiratory and gastrointestinal infections in particular younger children (63, 64). Following its first identification in Sweden, HBoV has been frequently detected in patients with various diseases throughout the Europe (65-67), North (68, 69) and South America (70, 71), Australia (72, 73), Asia (74, 75), and Africa (76, 77). In addition to respiratory and feces specimens, HBoV has also been identified in blood (78), urine (79), saliva (80), cerebrospinal fluid (81), and even environmental samples (82, 83). The current evidence clearly illustrates the potential risk of HBoV exposure for human health. In this review and meta-analysis, we summarized the results of 36 studies, including a total of 23,664 pediatric cases of gastroenteritis. The updated overall estimate suggested that HBoV infection is associated with 9.1% of all gastroenteritis cases in children, which confirms and extends the earlier results of a systematic review and meta-analysis of HBoV conducted by Ri De (22). This growing estimate emphasized that HBoV infection contributes to the increased global burden of gastroenteritis in children.

In comparison to the previous meta-analysis (22) that only involved children aged under five years, the age distribution of HBoV-associated gastroenteritis in children younger than 18 years was available in the present work. These data revealed that children under five years of age were the most susceptible to develop gastroenteritis when infected with HBoV, with the highest prevalence of HBoV-associated gastroenteritis cases (12.1%). From the results of more specific age groups, the relatively high frequencies of HBoV were also found in the age groups less than one year old (7.3%) and between 1 and 5 years old (5.2%). These findings tie well with previous reports wherein younger children might undergo a higher occurrence of HBoV-associated gastroenteritis. For example, a recent large-scale survey conducted in South Africa with 3765 fecal samples collected from hospitalized children for gastroenteritis illustrated that more than 90% of HBoV-associated cases were children younger than two years (84). A case-control study in the UK concluded that although HBoV is relatively common across all age groups, the risk of HBoV-associated gastroenteritis derived from preschool-age children was expected to be higher compared with adults (37). Furthermore, Mehrdad et al. reported that the significant age category in HBoV-positive samples was between 1 and 2 years old and have concluded that the prevalence of gastrointestinal tract infection with HBoV occurs precisely in the age range of digestive illnesses resulted from rotavirus (85). Our findings also matched several reviews focused on global norovirus (NoV) prevalence, demonstrating that most pediatric NoV-associated gastroenteritis occurred in children under two years of age (86, 87). Taken together, childhood gastroenteritis caused by HBoV and other known gastroenteritis-related viruses occurs precisely within the same age range. Thus, HBoV is much likely to figure as a potential etiological agent of gastroenteritis.

Comparisons across different countries showed that the highest HBoV prevalence in children with gastroenteritis was observed in Egypt (57.8%), followed by Australia (29.0%), Latvia (25.3%), and Brazil (24.7%). The lowest HBoV prevalence occurred in pediatric patients from Korea (0.7%). In line with the previous demonstration, these results confirmed that HBoV had been identified worldwide without any regional or geographic restrictions (63). However, these findings go beyond previous reports, which indicated a trend of decreasing prevalence from the developing countries to the developed countries (88, 89). For illustration, a case-control assessment conducted in the United States revealed that HBoV infection was related to 12.4% of all cases of acute childhood diarrhea (29). Nonetheless, the pooled HBoV prevalence was estimated at 5.8% among 1196 pediatric cases in India. It remains unclear to what complicated reasons attributed to the wide variation of HBoV prevalence across the world in addition to ethnicity, nutritional status, hygiene conditions, healthcare settings, and the various sample size of the study. More comprehensive studies should take into account the concrete reasons for HBoV prevalence in multiple geographies.

Consistent with previous evidence indicating less susceptibility to viral infections in females (90), our data supported that males are more susceptible than females to develop HBoV-associated gastroenteritis, with 9.4% and 8.7% of HBoV prevalence in male patients and female patients, respectively. However, no significant differences were observed in HBoV prevalence based on gender. Further research is needed to address this information lacuna.

Four HBoV genotypes have been described based on the genomic analyses of VP1 and VP2 sequences. Globally, HBoV1 accounted for more than half of all infections in children with gastroenteritis, which also dominated in most countries. It was also found that only one genotype was presented in Korea (HBoV1), Gabon (HBoV1), and Bangladesh (HBoV2). The estimated genotype distribution by country displayed considerable regional variations. As already mentioned, HBoV1 is commonly prevalent in clinical specimens of the respiratory tract, whereas HBoV2-4 are more frequently detected in patients with gastrointestinal infections and symptoms (64). Additionally, a recent relevant study in Taiwan demonstrated that HBoV1 and HBoV2 are generally the primary species in childhood gastrointestinal infections worldwide (43). The prevalence of HBoV genotypes has been investigated in children with gastroenteritis in the previous meta-analysis (22). The researchers showed that HBoV1 and HBoV2 were detected in 3.49% and 8.59% of pediatric cases with gastroenteritis. These previous results from Ri De et al. seem to be inappropriate for comparison because of which estimated data based on only ten case-control studies. In our updated review, HBoV1 (3.8%) figured as the predominant genotype circulating in children with gastroenteritis, and HBoV2, with a prevalence of 2.4% in the next rank. Even though HBoV1 can also be found in fecal samples from pediatric patients suffering from gastroenteritis, this novel finding suggested the prevalent HBoV variant changed related to childhood gastroenteritis during the past years. Likewise, during a 3-year observation in Russia, all four HBoV genotypes with differing detection rates have been identified in children hospitalized with diarrhea (91). Hence, we believe that continuous surveillance of this virus would be vital in pointing out how the prevalent HBoV genetic variants change with time. With regard to other genotypes, we identified the relatively low prevalence of HBoV3 in children with gastroenteritis (0.9%), while HBoV4 was only determined in four included studies. Since the lower prevalence and frequency of the other genotypes, it remains to be resolved whether other genotypes display a pathogenic role in childhood gastroenteritis.

Using data from 11 case-control studies, a meta-analysis provided new evidence to construct pooled risk effects of HBoV-associated gastroenteritis. It was found that children infected with HBoV have approximately 1.6 times increased chance of developing gastrointestinal symptoms compared with uninfected controls, indicating that HBoV exposure is significantly associated with a higher risk of gastroenteritis in children (OR 1.620, 95% CI: 1.023-2.566). Additionally, the presence specifically of all four HBoV genotypes was examined for the purpose of determining any possible associations between specific HBoV genotypes and gastroenteritis in children. The analytical results indicated that HBoV2 was significantly correlated with gastroenteritis in children (OR 1.746, 95% CI: 1.296-2.351), whereas other genotypes have not yet disease association. This supports Arthur and coworkers who have proven the association between HBoV2 infection and gastroenteritis using multivariate conditional logistic regression analysis (11). However, the pathogenic role of HBoV2 has been challenged by Ong et al. (92), demonstrating that HBoV in stool is probably an innocent bystander rather than a true pathogen due to high coinfection with other pathogens. It has also been questioned owing to the fact that HBoV was frequently detected in healthy or diarrhea-free individuals (17). In this updated review, the pooled prevalence of HBoV infection was estimated at 4.0% in children without gastroenteritis, which significantly lower than that in pediatric patients (*P* < 0.001). The most likely explanation for the presence of HBoV in feces from asymptomatic individuals can be linked to the long-term viral shedding following symptomatic infection (93, 94). This regard does not repudiate the role of HBoV as the etiological agent of gastrointestinal symptoms in children. Furthermore, the significantly positive association of HBoV infection with childhood gastroenteritis were observed in India (OR 26.34, 95% CI: 1.62-429.32), China (OR 1.70, 95% CI: 1.18-2.43), and Australia (OR 2.13, 95% CI: 1.29-3.52), while it was shown inverse association in the UK (OR 0.66, 95% CI: 0.51-0.86). The findings varied across countries might be due to the regional differences in viral epidemiology, geographic location, and seasonality (18, 95, 96).

Several limitations of the previous study (22) were addressed in the present work. However, it remains some limitations such as the lack of a large number of eligible research data and not considering other factors that might affect the results. Furthermore, the present work only included children diagnosed with gastroenteritis or diarrhea as cases, making it probably underestimate the burden of HBoV infection on the development of childhood gastroenteritis.

## Conclusion

In summary, the pooled prevalence of HBoV infection in children suffering from gastroenteritis was estimated at 9.1% between 2016 and 2021, showing a higher global burden of gastroenteritis in children caused by HBoV than in the last decade. The considerably higher HBoV prevalence was found in pediatric patients less than five years of age and in Egypt. Notably, the correlation of HBoV infection with gastroenteritis in children was established in this updated review. Our findings also add to the growing evidence that HBoV2 would be a causative agent of gastrointestinal symptoms. However, this conclusion should be treated with caution as the known gastroenteritis-associated viruses are not excluded. The actual pathogenic role of HBoV in leading to childhood gastroenteritis were needed to be determined by large-scale continuous surveillance, serological investigation, and appropriate animal models.

## Data Availability

All data produced in the present work are contained in the manuscript

